# Person-to-Person Microbiota Transmission Can Influence Depression and Anxiety in Newly Married Couples: Six-Month Interim Results

**DOI:** 10.1101/2025.02.07.25321564

**Authors:** Reza Rastmanesh, Balachandar Vellingiri, Ciro Gargiulo Isacco, Abolfazl Sadeghinejad, Neil Daghnall

**Author notes:** Corresponding author: Reza Rastmanesh, # 6 Physicians Building, Sarshar Alley, Tajrish, Tehran, Iran, Postal Code: 1961835555.

## Abstract

**Background:** Oral microbiota dysbiosis and salivary cortisol are associated with depression and anxiety. Bacterial transmission can occur between spouses.

**Aims:** We explored whether oral microbiota, salivary cortisol and a combined depression-anxiety (DA) phenotype affiliated in newly married couples.

**Methods:** The researchers administered validated Persian versions of the Pittsburgh Sleep Quality Inventory (PSQI), Beck Depression Inventory-II (BDI-II), and Beck Anxiety Inventory to 1740 couples, who had been married during the past six months. The investigators compared 296 healthy control spouses with 296 cases. Data analysis used appropriate statistical methods.

**Results:** After six months, at the phyla level, we identified a significant decrease in Firmicutes and Actinomycetota abundance and an increase in Bacteroidetes, Proteobacteria, Fusobacteria and Patescibacteria abundance in healthy spouses married to an insomniac with DA-phenotype, showing that oral microbiota were significantly changed and became similar to that of participant’s spouse, (i.e., if the spouse had DA-phenotype, then composition of oral microbiota became similar to their spouse DA-phenotype, *p* <0.001). These changes parallelled alterations in salivary cortisol, depression and anxiety scores. Linear discriminant analysis (LDA) showed that relative abundances of Clostridia, Veillonella, Bacillus and Lachnospiraceae were significantly higher in insomniacs with DA-phenotype than healthy controls (*p* <0.001). Results remained significant after controlling for confounders. The formal mediation analysis confirmed these outcomes. We observed distinct analogous gender differences for oral microbiota pattern, salivary cortisol level, and depression and anxiety scores.

**Conclusions:** Microbiota transamination between two people in close contact with one another partially mediated depression and anxiety.

## INTRODUCTION

Oral microbiota dysbiosis correlates significantly with multiple neuropsychiatric diseases such as autism spectrum disorder, dementia, Parkinson’s disease, schizophrenia, anxiety, epilepsy and depression ^1-2^. Wingfield et al in a preliminary study showed that oral microbiota composition is significantly associated with depression in young adults who met the DSM-IV criteria for depression. Overall, twenty-one bacterial taxa were significantly differentially abundant in the depressed young adults, including increased Prevotella nigrescens and Neisseria spp., whereas nigteen taxa had a decreased abundance ^3^.

Oral microbiome involving enhanced salivary cortisol and pro-inflammatory communication, may mediate oral health and mental health ^4-5^. Simpson et al recently showed that oral microbiota composition is correlates significantly with anxiety and depression symptoms in adolescents ^6^. Similar patterns have been noted in other groups, such as pregnant women ^7^, patients suffering from burning mouth syndrome with psychiatric symptoms ^8^, patients with anxiety, mood and trauma- and stress-related disorders ^9^ and irritable bowel syndrome ^10^, just to mention a few.

The first author has elsewhere ^11^ discussed how closeness may facilitate bacterial transmission and consequently, impact microbiota-related research. Prior research has reported other types of physiologic synchrony between couples. For instance, synchrony of diurnal cortisol pattern ^2^, cardiac synchrony ^12^ and sleep concordance ^13^. The bi-directional associations between sleep disturbances ^14^ and ocular surface parameters ^15^ with oral/gut microbiota compositions, along with other physiological synchronies ^2, 12-13^ between couples lead the authors to hypothesize that oral microbiota transmission partially mediated depression. Finally, it is known that cortisol is a biomarker of anxiety and depressive states in couples and partners ^16-17^.

Based on these interconnected premises, we hypothesized that oral microbiota partially mediated psychometric parameters in newly married couples and that this occurred via person-to-person contact. To assess this novel supposition, the researchers enrolled couples in whom one spouse simultaneously suffered from depression and insomnia (hereafter: DA-phenotype). To facilitate contact spouses lived in a same house.

## Methods

### Participants

Data were collected prospectively from two private sleep clinics, Tehran, Iran.

### Screening for insomnia, depression and anxiety

Participants who had been married during the past six months and were in a cohabiting relationship were screened based on having or not having insomnia. Those selected were enrolled to participate in the study together with their official spouses. Participants then were categorized to (i) healthy and (ii) insomniac spouses. Each group were further divided by BDI-II and BAI scores into “moderate depression” and “moderate anxiety”. A combined depression-anxiety phenotype (DA-phenotype) in this study was defined as simultaneously having BAI: 16-25 and BDI-II ≥14.

296 couples were selected as healthy spouses and insomniacs with DA-phenotype. Three couples were excluded because the women were taking medicines known to affect oral microbiota composition or were pregnant. No female spouses used medications known to interact with the hypothalamic-pituitary-adrenal axis. Seventeen were excluded due to either low readings (n = 10) or missing data (n = 7). One couple divorced from each other and had spent significant time living apart and seven couples who had moved to another city were excluded.

The remaining 268 couples lived with their spouses in a same house. On Day 1 and Day 180, all couples participated in oral microbiota composition and salivary cortisol measurement study. Data drawn from 268 couples were analyzed. The diagram of categorization and enrollment of participants is shown in **Figure 1**.

**Figure 1.**
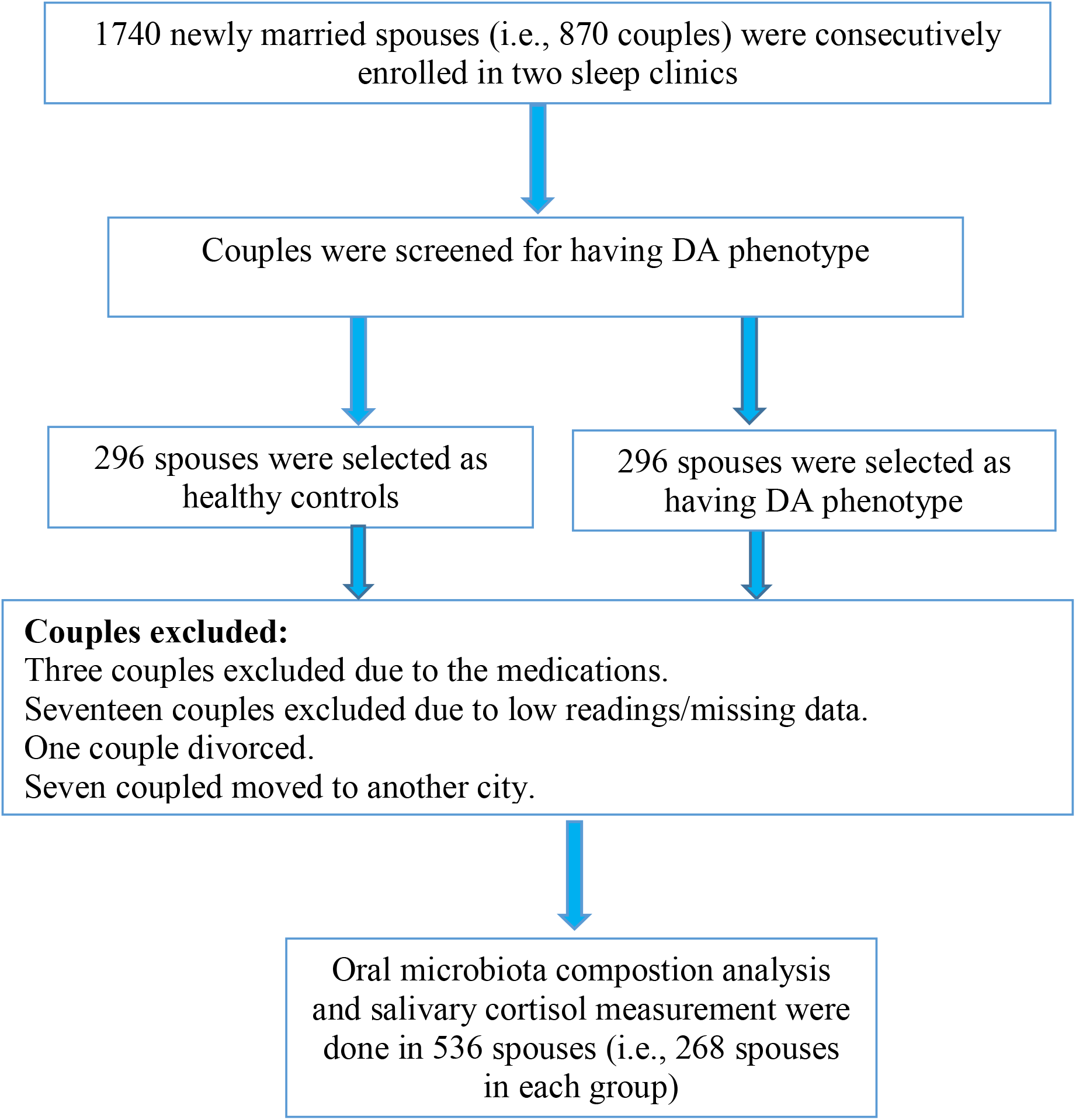
Diagram of categorization of participants according to sleep disturbance status and depression-anxiety (DA) phenotype

**Figure 1.**
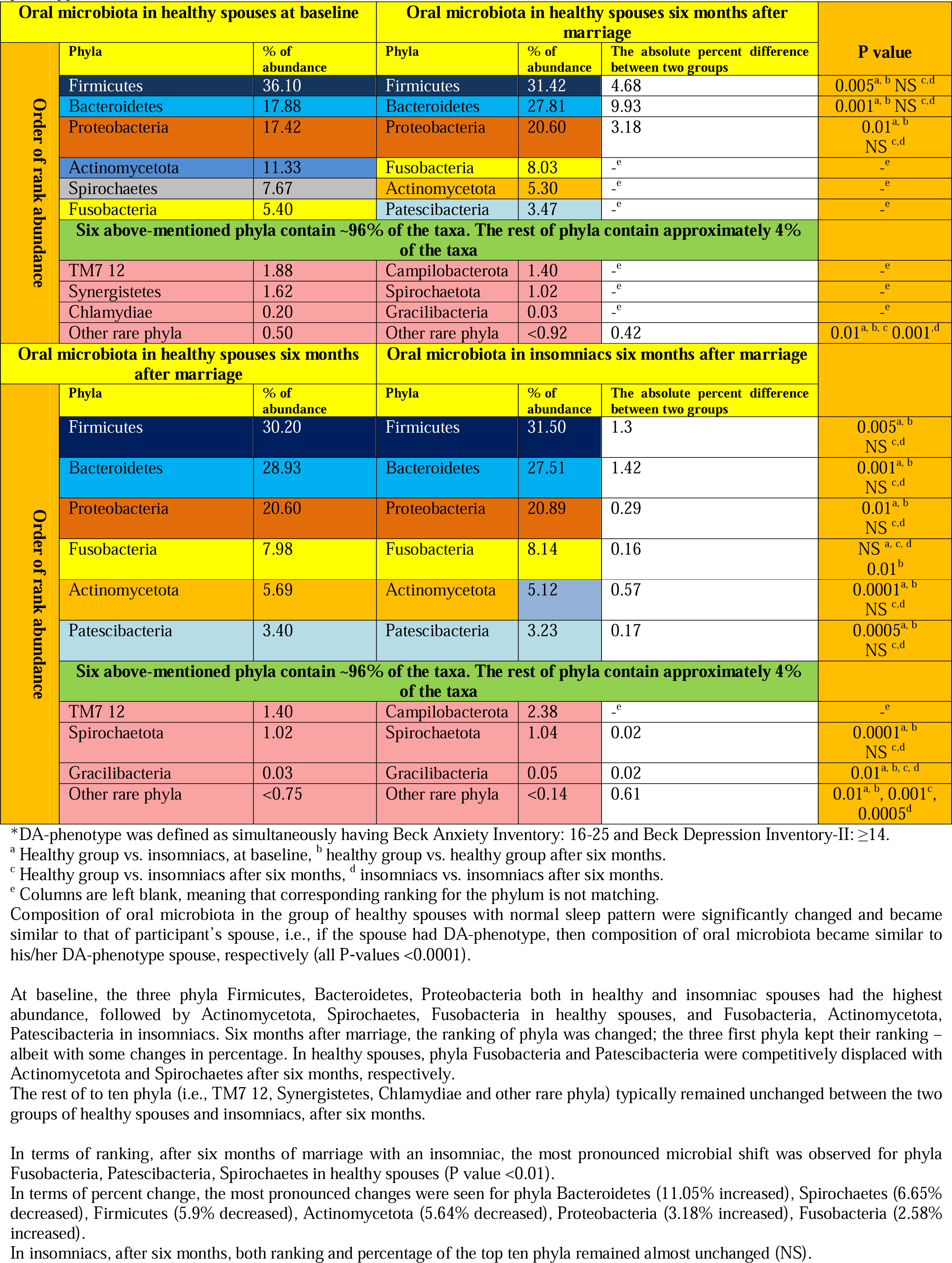
Baseline and six-month comparison of oral microbiota in healthy spouses with spouses who had depression-anxiety phenotype*

## Research instruments

### Psychometric assesments

The validated Persian version Beck Depression Inventory-II (BDI-II) ^18^, Beck Anxiety Inventory (BAI) ^19^, and Pittsburgh Sleep Quality Index (PSQI) ^20^ were employed to measure depression, anxiety, and sleep quality, respectively.

The BDI-II is a commonly utilized 21-item self-report inventory designed for assessing depressive symptomatology, referring to the last two weeks. Higher scores demonstrate more severe depressive symptomatology. Depression in this study was defined as a BDI-II score ≥14 ^21^.

The self-administered PSQI questionnaire measures the sleep quality over the past. The scores range from 0-21; a lower score represents better sleep quality. Scores higher than 5 is regarded poor sleep quality. Insomnia (sleeplessness) was defined as coexistence of both difficulty resuming sleep and daytime dysfunction ^22^.

BAI questionnaire has 21 items with scores ranging from 0-3 for each item, in which 0 indicates “not at all,” 1 indicates “mildly, but it did not bother me much,” 2 indicates “moderately, it was not pleasant at times,” and 3 indicates “severely, it bothered me a lot” for all the items. The total score ranges from 0-63, which demonstrates levels of anxiety. Scores from 0-7 indicates a minimal range (no anxiety). Scores of 8-15 shows mild, 16-25 indicates a moderate range, and Scores of ≥26 represents severe anxiety. Additionally, in comparison with category of no anxiety, the three categories of mild, moderate, and severe anxiety were considered as “yes” ^23^.

We did not include any severe symptomatic spouses who would have needed additional psychological care.

### Oral parameters

Oral samples were collected at clinic visits from enrollment until 5-6 months after marriage, as briefly described below:

#### Oral microbiota

We followed the guidelines developed by Brzychczy-Sroka et al ^24^. Accordingly, we collected the oral samples from the palatine tonsils at the end of the examination before posterior pharynx swab collection. Samples were secured in saline, placed in ziplock bags, secured in portable freezer bags at − 20 °C, and then delivered immediately to the laboratory (maximum 1-2 h).

Samples were then frozen to −80 °C untill analyses. One of the researchers (RR) secured collected materials in the MoBio buffer and placed them in a small ice-flled cooler for transport without defining a precise transport temperature, exactly as suggested by Brzychczy-Sroka et al ^24^. Final materials were delivered to the laboratory within 3-4 hours.

### Salivary cortisol

Salivary samples were collected from particiants using Oragene OG-500 kits (DNA Genotek, Ontario Canada), that enables self-collection and stabilisation of DNA at room temperature. Couples were instructed not to consume any food or drink, except water, for at least 30 min prior to sample collection. Saliva cortisol was then measured by LC-MS/MS (as described previously) ^25^. To avoid risk of drift in the analytical method, saliva specimens were stored at −80 °C for concomitant analysis. We used samples provided immediately on awakening before drinking, eating, or doing oral hygiene.

### Statistical Analysis

Statistical analyses were performed with SPSS software (ver. 17.0; SPSS, Chicago, IL). Differences between groups and subgroups were analyzed by Student’s t-test for continuous parameters and the χ^2^ test for categorical parameters. Intragroup changes were compared with paired t-test. Primary and secondary endpoints were analzyed using analysis of covariance with groups as fixed factors and baseline measurements as covariates. Furthermore, Pearson’s and Spearman’s correlation tests were applied to explore correlations between oral microbiota, anxiety depression, and insomnia. Oral microbiota analysis was done using QIIME 2 2019.07 ^26^. Processed data were imported into phyloseq v1.28.024 ^27^ for analysis. Alpha and Beta diversity was evaluated using Shannon’s diversity index and Bray-Curtis dissimilarity. For a detailed description on the methodology used for bioinformatic processing please refer to ^3^. To determine which taxa may be correlated with DA-phenotype, we performed L2-regularized logistic regression using the mikropml package R ^28^, which is a commonly applied methodology for performing differential abundance analysis of microbiota data ^29^.

### Covariates

Gender, age, body mass index (BMI), drinking and smoking status, total dietary sugar intake, and presence of chronic kidney disease (CKD) or high blood pressure were considreded as covariates. High blood pressure was defined as an average systolic blood pressure ≥140 mmHg and average diastolic blood pressure ≥90 mmHg, or use of antihypertensive drugs or a physician diagnosis or. CKD was defined as an eGFR <60 ml/min/1.73 m^2^.

## RESULTS

**Table 1** shows participant baseline characteristics. Demographic characteristics, such as gender and age, BMI, and socioeconomic status were similar between healthy controls and DA-phenotype group (**Table 1**). Mean ± SD for age in male and female spouses were 37.20 ±8.01 and 31.02 ±9.30 years, respectively. Couples had been married and living together for an average of 5.91 ± 2.03 months. As expected, there were significant differences for salivalry cortisol levels, global PSQI, BDI-II and BAI scores between the healthy controls and the insomniacs with DA-phenotype group at baseline.

### Psychometric parameters

**Table 2** shows that after six months, healthy spouses who had married to an insomniac spouse having DA-phenotype scored significantly higher on PSQI, BID-II and BAI compared to their own baseline values. This indicated sleep quality, depression and anxiety scores changed and became similar to that of participant’s insomniac spouse, but still was significantly lower than that of the inscomniacs group (*p*< 0.001). After six months, in insomniacs with DA-phenotype, there there was a trend of increased salivalry cortisol levels, global PSQI, BDI-II and BAI scores, but values did not reach significance (non-significant differences are not shown).

In gender subgroup analysis, worsening of insomnia severity, increment of salivary cortisol, and increase in depression and anxiety scores were more pronounced in female spouses after six months (**Table 2**).

Multiple logistic regression analysis demosntarted that female gender, higher level of education, older age, having lower income, residing near highway/street and cigarette smoking were associated with insomnia severity, depression, and anxiety scores.

In the crude model, insomnia severity was positively associated with anxiety and depression scores (OR: 1.62, 95% CI: 1.32–2.19, *p* < 0.001, OR: 2.30, 95% CI: 2.10–2.75, *p* < 0.001, respectively). After adjusting for age, gender, BMI, and education, insomnia remained positively correlated with depression and anxiety scores (showing a 1.8-fold and 2.1-fold, respectively) and higher odds compared to healthy spouses with normal sleep (OR: 1.8, 95% CI: 1.40–2.52, *p* < 0.001, and OR: 2.1, 95% CI: 2.01–2.73, *p* < 0.001).

### Salivary cortisol differences

**Table 1** shows t-test analysis of salivary cortisol. In both male and female spouses with DA-phenotype, salivary cortisol was significantly higher in insomniac spouses at baseline compared to the healthy control spouses (*p* < 0.0001). After six months, salivary cortisol level in spouses who had married to an insmoniac DA-phenotype spouse were significantly increased compared to baseline values (*p* < 0.0001). This suggested that healthy spouses were likely to resemble their insomniac spouses who had DA-phenotype. Gender subgroup analysis showed that the increased salivary cortisol levels in female spouses were more noticeable than male spouses (**Table 2**).

### Oral microbiota characteristics

From the view of taxa composition, a total of 33 bacterial phyla were identified. Linear discriminant analysis (LDA) of oral microbiata composition showed that the relative abundances of Clostridia, Veillonella, Bacillus and Lachnospiraceae were signicantly higher in the DA-phenotype spouses than healthy controls (*p* < 0.001, LDA scores > 2, alpha error = 0.01).

A high-level view of phyla differences found that the composition of oral microbiota in healthy spouses with normal sleep pattern were significantly changed and became similar to that of participant’s spouse (i.e., if the spouse had DA-phenotype, then composition of oral microbiota became similar to his/her DA-phenotype spouse, respectively *p* < 0.001, LDA scores > 2, alpha error = 0.01) (**Figure 2**).

The relative abundance of Fusobacteria (*r* = +0.49–0.57), Patescibacteria (*r* = +0.38–0.42), Campilobacterota (*r* = +0.32–0.36), Spirochaetota (*r* = +0.42–0.52), and Gracilibacteria (*r* = +0.29– 0.37) in the oral microbiota were positively correlated with severity of insomnia in insomniacs with DA-phenotype (all *p* values < 0.01).

We applied a L2-regularized logistic regression model on the next generation sequencing data to investigate potential associations between oral microbiome and DA-phenotpe. This model showed that Fusobacteria, Patescibacteria, Campilobacterota, Spirochaetota, and Gracilibacteria may be connected with DA-phenotype.

### Gender differences

LDA and multivariable association analysis showed multiple distinct, significantly differently abundant bacterial genus between female and male spouses who had DA-phenotype (*p* < 0.001, LDA scores > 2, alpha error = 0.05). Phylum Proteobacteria was significantly more abundant in females sposues with DA-phenotype than in male sposues. Also, some members of phyla Firmicutes and Bacteroidetes were significantly more abundant in these female spouses. Intrestingly, genus Dialister (family Firmicutes) was significantly more abundant in female spouses with DA-phenotype than in male spouses (all *p* values < 0.001).

### Mediation Analysis

To better understand the role of oral microbiota in salivary cortisol and DA-phenotype status, we performed interaction effects assessment and mediation analyses. We suspected interaction effects between these three variables with oral microbiota status as the putative mediator of the link between salivary cortisol and DA-phenotype status. In fact, oral microbiota pattern is a significant predictor of DA-phenotype status (β = 0.37, *p* < 0.001) which in turn is also a relevant predictor for salivary cortisol (β = 0.15, *p* < 0.001). The mediation analysis explained 24% of the variability in the data (R^2^= 0.24).

## DISCUSSION

Oral microbiota transamination between two people in close contact with one another (i.e., the couples in the present study mediated depression and anxiety. Although there is no identical human study to compare these findings with, there are many evidence of bacterial exchange between humans with dogs ^30^, chimpanzees ^31^, and Livestock ^32^. There are also reports on penile/genital microbiota exchange between partners ^33-34^. These finding underlie the importance of bacterial exchange as a potential mediating mechanism of mood synchrony between spouses and partners. Many other types of physiological synchrony between spouses and partners has been reported before, such as cardiac synchrony ^12^, diurnal cortisol pattern synchrony ^2^, and also sleep concordance ^7, 13, 35^.

We showed that changes in oral microbiota composition is associated with changes in severity of insomnia ^7, 36-38^, salivary cortisol levels ^6, 9^, and depression ^2-10^ and anxiety ^2-10^ scores. Our finding is in accordance with previous studies ^2-6, 8-10, 36-38^.

Recently, in a large cohort of couples who had been married and living together for an average of 5.91 months, we showed that sleep disturbances can be partially explained by changes in gut microbiota ^14^. Also, in another study we discovered a significant association between ocular microbiota and dry eye disease (DED) in insomniacs, which was possibly mediated through person-to-person contact ^15^. In that study, we showed that six months after marriage, spouses who were married to an insomniac with DED phenotype, were more significantly likely to develop DED after six months follow up. In support of our initial hypothesis, these changes occurred in parallel to the changes in ocular microbiota composition ^15^.

Oral–gut microbiome are connected to each other ^39-40^, with distinct similarities and differences ^40^. On the other hand, there are known connections between oral microbiota and ocular microbiota ^41-44^. Furthermore, salivary cortisol is a predictor of depression ^45^, anxiety ^46^, insomnia ^47^ and DED ^48-49^. Overall, this suggests that these networks interconnect. Indeed, current literature supports such a conclusion. The frequency/prevalence of DED in people who are depressed or have anxiety is significantly higher than healthy people and vice versa ^7, 50-53^. Therefore, our findings have implications for holistic medicine, family medicine and personalized medicine. The practical and theoretical implications of this study cover vast areas of sleep therapy and psychological states.

This research was a non-invasive prospective observational study in nature, however, there are preclinical convincing evidence to support our hypothesis; transplantation of fecal microbiota from patients with depression to microbiota-depleted rats can induce physiological and behavioral properties characteristic of depression in the recipient animals ^54^. Furthermore, Lee et al showed that fecal microbiota-induced insomnia, immobilization stress and depression-like behaviors in mice model, is alleviated by microbiota-modulating probiotics ^55^. These two last reports ^54-55^ strongly suggest the existence of a causal association between microbiota changes and psychometric parameters such as depressive states and anxiety. Taken together, our preliminary findings provide evidence that the association between oral microbiota changes and mood changes or its synchrony in humans are causally related ^55^. Future research should further evaluate this supposition.

### Strength

Firstly, this study employed a robust methodology, specifically a large sample size for an acceptable follow up program. Secondly, to our knowledge, this is the first study of its type to investigate a large array of variables together. Thirdly, although not homogeneous, exclusively selecting and recruiting this type of participants enabled us to study effect of continuous and short-midterm oral bacterial transmission on depression, anxiety in a real-word situation. We used a combined depression/anxiety scale, which is more informative than depression and anxiety scores alone.+44

### Limitations

After six months, worsening of insomnia severity, depression and anxiety scores was more noticeable in female spouses. It is possible that we have underestimated/under-reported the frequency of insomnia, depression and anxiety in male spouses, because we used PSQI, BDA-II and BAI to estimate insomnia, depressive and anxiety symptoms, respectively. It is important to acknowledge that our participant couples were aware of the purpose of the study, due to the ethical constraints. A main effect of condition has been found in previous well-controlled and well-conducted researches such that both males and females reported significantly more insomnia severity ^56-57^, depression ^21, 58^ and anxiety symptoms ^59-60^ in the covert condition, suggesting that we may have underestimated/under-reported the frequency of insomnia, depression and anxiety in males spouses while have overestimated/over-reported these parameter in female spouses. Despite controlling for the most important covariates, additional residual confounding may exist. For instance, dietary food intake is animportant factor influencing the composition of the gut microbiome. Dietary Index for Gut Microbiota (DI-GM) ^61^ has been suggested to account for this covariate. However, this tool is not mature enought at the moment to enhance the precision of designs. Addressing these issues is a topic for future work. Couples were not randomly selected, therefore the results are not completely generalizable to every circumstances. Finally, we only measured morning salivary cortisol, and measurement of salivay cortisol in three consecutive days could enhance accuracy. Finally, we only used BDI-III and BAI, while clinical diagnosis of anxiety and depression based on DSM is a more precise method ^62^.

## Contributors

RR, CGI and ND designed this research. BV and AS developed the concept for the manuscript. RR and AS coordinated the implementation of this research and contributed to providing medical data. RR and BV wrote the manuscript. RR and CGI contributed to the analysis of this research. All authors have read and approved the final manuscript. RR is responsible for the overall content (as guarantor).

## Patient consent for publication

Consent obtained from participants.

## Ethics approval

The study was approved by the ethics committee of the Iran National Science Foundation (INSF) (Research Ethical Code INSF: 98R026323-2024) and adhered to the tenets of the Declaration of Helsinki. Participants gave informed consent to participate in the study before taking part.

## Data availability statement

Data are available on reasonable request.

## Acknowledgenemts

We would like to thank the couples who participated in this study. We also wish to especially thank Dr. Javid Azizi very much for his generous financial support of this project, and for his careful involvement in each stage of the work. Finally, we would like to thank to the managers of private sleep clinics for their brilliant cooperation.

